# Long-Term Slowing of Progression in Huntington’s Disease with Pridopidine Treatment

**DOI:** 10.64898/2026.02.13.26345490

**Authors:** Ralf Reilmann, Andrew M. Tan, Anne E. Rosser, Kelly Chen, Karen E. Anderson, Sandra K. Kostyk, Andrew Feigin, Randal Hand, Michal Geva, Michael R. Hayden

## Abstract

**Background:** Huntington’s disease (HD) causes progressive loss of function, cognition, and motor control, with no approved therapy yet shown to slow disease progression. In the PROOF-HD phase 3 trial, pridopidine did not meet the primary or key secondary outcomes in the overall population, but participants who remained off antidopaminergic medications (ADMs) showed benefits compared to placebo during the double-blind phase. Whether such benefits continue with longer duration treatment and how they compare with expected natural-history trajectories remains unknown.

**Methods:** We evaluated outcomes through Week 104 from baseline in participants who received continuous pridopidine (45 mg twice daily) and remained off-ADMs throughout the double-blind and open-label extension period (n=90). External comparators from ENROLL-HD and TRACK-HD were constructed using propensity-score weighting methods. Least-squares mean changes from baseline to Week 104 were estimated using mixed-effects models for repeated measures across outcomes.

**Results:** At two-years, pridopidine treatment was associated with benefits versus propensity-score weighted natural-history comparators across multiple outcomes. Relative to ENROLL-HD, participants receiving pridopidine showed slowing of progression over 104 weeks, expressed as percent slowing across cUHDRS, TFC, SWR, SDMT, and TMS outcomes (39.5–88.3% slowing). Similar patterns were observed relative to TRACK-HD across the same measures (48.5 – 81.5% slowing), including quantitative motor performance assessed by Q-Motor FT-IOI (77.8% slowing). Exploratory analyses including participants receiving concomitant ADMs showed similar directional patterns as the primary analyses.

**Conclusions:** In a two-year follow-up, continuous pridopidine treatment in participants remaining off-ADMs was associated with slower clinical progression relative to expected natural-history trajectories.

(Clinical Trials Identifier: NCT04556656)

## Introduction

Huntington’s disease (HD) is a rare, progressive neurodegenerative disorder that typically progresses over 10–20 years and is characterized by declining functional capacity, cognition, and motor control ^1–3^. In manifest HD, clinical progression is quantified using standardized outcomes that capture function, cognition, and motor impairment, including the Total Functional Capacity (TFC) score, composite Unified Huntington’s Disease Rating Scale (cUHDRS), Stroop Word Reading (SWR), Symbol Digit Modalities Test (SDMT), and Total Motor Score (TMS)^4–7^. Worsening scores over time reflect increasing disease burden and loss of independence^2,8^.

Despite a well-characterized natural history and the substantial toll of HD on individuals and their caregivers, no approved therapy has been shown to slow disease progression. Pridopidine is a selective sigma-1 receptor (S1R) agonist in late-stage clinical development in HD and ALS ^9–11^. In the global phase 3 PROOF-HD randomized, double-blind, placebo-controlled trial (NCT04556656), pridopidine 45 mg twice daily did not significantly differ from placebo on the primary or key secondary endpoints at Week 65 in the overall population ^10^. However, participants who remained off-ADMs throughout the double-blind phase (DBP) showed differences favoring pridopidine across multiple functional, cognitive, and motor outcomes, and measures of progression^10^. Whether such observations persist with longer term follow-up, and how they compare with expected trajectories in untreated early HD is clinically very relevant and important ^2,8,12,13^. We therefore examined whether longer-term trajectories with continuous pridopidine treatment showed consistent slower progression across clinical outcomes relative to expected natural-history disease trajectories estimated from external cohorts.

The PROOF-HD protocol included an open-label extension to evaluate active treatment from the original DBP baseline through Week 104^10^. In the absence of a placebo group, trajectories were interpreted using external natural-history comparators from ENROLL-HD and TRACK-HD, which provide robust longitudinal data in early HD ^6,12,14,15^. To improve baseline comparability, we applied well-established propensity-score (PS) methods with trimming and Average Treatment Effect on the Treated (ATT)-oriented weighting^16–19^. While recognizing the limitations inherent to an open-label, externally controlled design^19^, this approach enabled clinically interpretable comparisons of longer-term disease progression in settings where prolonged placebo control is no longer feasible.

Prior work has shown that relatively small numerical changes in the cUHDRS and its sub-component measures can correspond to meaningful differences in daily functioning and, if sustained, may preserve independence over time ^4,7,13,20^. As proposed by Schobel et al., a slowing of cUHDRS decline corresponding to a ∼20–30% reduction in the annual rate of progression has been suggested as clinically meaningful ^7^. To support clinical interpretation, outcomes were summarized using least-squares mean changes from baseline, between-group differences as compared with PS–weighted natural-history comparators, and descriptive estimates of percent slowing of progression. These summaries were intended to support patient-relevant interpretation of treatment-associated differences, while recognizing the inherent limitations of an open-label, externally controlled design for inferring long-term treatment effects on disease trajectory^10,13,15–17^.

To assess robustness under routine clinical conditions, exploratory analyses incorporated concomitant antidopaminergic medications (ADMs) to examine whether pridopidine treatment-associated trajectories persisted with ADM use. Although ADMs provide symptomatic benefit, ADMs may also adversely influence longitudinal measures of trial outcomes in HD ^15,21–25^. Together, this study evaluated whether continuous pridopidine treatment was associated with slower clinical progression in early HD as compared with expected natural-history progression.

### Objective Statement

The objective of this study was to determine whether continuous treatment with pridopidine is associated with slower progression in early HD as compared with expected progression estimated from propensity-score weighted natural-history cohorts.

## Methods

### Study Design

PROOF-HD was a phase 3, randomized, double-blind, placebo-controlled study followed by a prespecified open-label extension (NCT04556656). The primary endpoint was assessed at Week 65. After completing the DBP, participants could enter a planned open-label extension (OLE) to evaluate long-term outcomes under continuous pridopidine exposure. For analyses presented here, outcomes were evaluated from the randomized DBP baseline through Week 104.

All participants entering the follow-up received open-label pridopidine 45 mg twice daily. Randomized DBP treatment assignments remained undisclosed to participants, investigators, and sponsor personnel throughout the follow-up period, until OLE database lock (May 24, 2024), to limit bias related to knowledge of prior randomized assignment. The SAP prespecified use of natural-history external comparators from ENROLL-HD and TRACK-HD, constructed using propensity-score weighting and prespecified visit mapping framework aligned to the PROOF-HD DBP baseline. SAP addenda prespecified off-ADM analysis populations for long-term analyses. The analytic population was defined as participants who remained off-ADMs throughout the DBP and OLE. Additional methodological details, including SAP addenda definitions, visit-window specifications, external comparator construction, and sensitivity analyses, are provided in the **Supplementary Appendix.**

The protocol and all amendments were approved by institutional review boards or independent ethics committees at each participating site. The trial was conducted according to the principles of the Declaration of Helsinki and the International Council for Harmonisation Good Clinical Practice guidelines. Written informed consent was obtained from all participants at initial enrollment, with reconsent required for participation in the long-term follow-up.

### Participants and Analysis Populations

Participants eligible for inclusion in the long-term continuous-treatment analysis were those who completed the End-of-Study visit of the double-blind period (DBP) without major protocol deviations and who proceeded into the open-label extension. All participants initiating the extension entered a two-week titration phase of pridopidine 45 mg once daily before receiving the open-label target dose of 45 mg twice daily.

To minimize confounding arising from concomitant medications, the primary analytic population for DBP portion was restricted to participants who were off-ADMs during this period. ADMs, including VMAT2 inhibitors and dopamine D receptor–acting neuroleptics, may confound pridopidine-associated effects or worsen measures of clinical outcomes, and therefore ADM exposure was explicitly delineated in the SAP^10,25^. The off-ADM group consisted of participants who did not receive any ADMs at any time during the DBP or the OLE. The primary analytic population included participants who remained off-ADMs throughout the DBP and open-label extension and contributed follow-up data through Week 104.

A secondary subgroup consisted of participants receiving recommended doses of selected ADMs. Based on SAP definitions, the recommended lower-dose thresholds were tetrabenazine ≤50 mg/day, deutetrabenazine ≤24 mg/day, aripiprazole ≤5 mg/day, and quetiapine ≤50 mg/day (see **Supplemental Table 3**). Participants receiving more than one ADM concurrently or any ADM other than the four agents permitted at recommended doses were excluded, as high-affinity D antagonism may confound functional, motor, cognitive outcomes and measures of progression ^25–29^.

### External Comparator Cohorts and Statistical Analysis

ENROLL-HD is a large, multinational, prospective natural-history registry that includes more than 20,000 participants across 23 countries and provides annual assessments of functional, cognitive, and motor outcomes using instruments identical to those in PROOF-HD ^12,14^. TRACK-HD was a prospective, multicenter longitudinal study of premanifest and early HD, providing deep clinical, imaging, and biomarker phenotyping through annual follow-up ^5,6^.

Efficacy analyses were performed within the off-ADM pridopidine continuous-treatment populations. The analysis population consisted of the modified intent-to-treat (mITT) cohort restricted to off-ADM participants during the DBP. For the OLE portion, only participants who entered the OLE and remained off -DMs were included. This cohort served as the treated group for construction of the ENROLL-HD and TRACK-HD external comparators and all Week-104 external-control analyses. The core outcomes required for comparison with PROOF-HD included cUHDRS, TFC, SWR, SDMT, and TMS. TRACK-HD additionally provided Q-Motor assessments, including Finger-Tapping Inter-Onset Interval (FT-IOI) Mean, enabling comparator analyses of objective motor function.

For external comparisons with natural history cohorts, follow-up periods in ENROLL-HD and TRACK-HD were re-anchored to the PROOF-HD double-blind baseline, with outcomes aligned to the nearest comparable annual visit intervals. Because both observational studies used annual assessment schedules, PROOF-HD follow-up to 52 and 104 weeks corresponded to Year 1 and Year 2 visits in the external cohorts. Visits in ENROLL-HD and TRACK-HD were assigned to Year 1, Year 2 analysis time points using prespecified day windows centered on ∼12 and ∼24 months after baseline. These windows were applied consistently across analyses to ensure comparable exposure durations between PROOF-HD and the external comparators. For Q-Motor outcomes, only TRACK-HD contributed data because ENROLL-HD did not collect Q-Motor assessments.

Comparator participants were selected to match the eligibility characteristics of the PROOF-HD follow-up population. For ENROLL-HD, inclusion criteria required age ≥25 years, a pathogenic CAG repeat length ≥36, Diagnostic Confidence Level (DCL) of 4,

TFC ≥7 (corresponding to early manifest stages 1–2), TMS ≥20, Independence Scale ≤90, and residence in North America or Europe. TRACK-HD participants were required to meet criteria consistent with manifest HD, defined by DCL-4 and functional and motor characteristics comparable to the PROOF-HD early manifest population. Because the PROOF-HD analyses focused on participants off-ADMs, external comparator participants were likewise required to be off-ADMs during the relevant assessment periods.

Separate propensity-score models were developed for ENROLL-HD and TRACK-HD under an ATT framework, using the PROOF-HD off-ADM analysis population as the treated group. Logistic regression was used to estimate the propensity score. Baseline covariates included age, sex, geographic region, CAG repeat length, baseline CAP100 (where available), and baseline clinical outcomes (TFC, cUHDRS, SWR, SDMT, and TMS); education level was included for ENROLL-HD.

ATT weights under inverse-probability weighting were applied to participants in the natural-history cohorts. Trimming thresholds prespecified in the SAP excluded individuals with propensity scores <0.05 or >0.95 to reduce extrapolation beyond the zone of empirical overlap. Standardized mean differences (SMD) were used to assess covariate balance before and after weighting.

Weighted MMRM analyses were used to estimate LS mean change from baseline through Week 104. Fixed effects included visit, baseline value of the outcome, HD stage, and region, with a random intercept for each participant. Residual covariance structure followed the hierarchy specified in the SAP. No imputation was performed; the model assumes data are missing at random. Exploratory ADM analyses applied the same MMRM framework within ADM-defined subgroups and, where external comparators were constructed, incorporated the same propensity-score weighting approach.

Differences in assessment schedules between the observational cohorts required outcome-specific decisions regarding comparator selection. TRACK-HD provided Q-Motor FT-IOI assessments, enabling comparison with PROOF-HD on objective motor outcomes, whereas ENROLL-HD did not include Q-Motor and therefore could not contribute to these analyses.

### Outcomes, Timepoints, and Interpretation Framework

Functional progression was assessed using TFC. Global clinical progression was evaluated with cUHDRS, which integrates TFC, TMS, SWR, and SDMT. Cognitive performance was evaluated with SWR and SDMT, motor function with TMS, and objective motor performance with Q-Motor FT-IOI (TRACK-HD only).

Analyses focused on change from DBP baseline to Week 104. PROOF-HD included scheduled assessments at Weeks 26, 39, 52, 65, 78, and 104, whereas external comparators contributed annual data at Year 1 and Year 2. Weighted MMRM incorporated all available PROOF-HD follow-up visits to estimate longitudinal trajectories; however, externally controlled comparisons are reported at Week 104 (Year 2) as the prespecified long-term timepoint, aligned with annual assessments in ENROLL-HD and TRACK-HD. Changes were referenced to DBP baseline to preserve the continuous-treatment framework.

Efficacy outcomes were summarized using least-squares (LS) mean change from baseline estimated from weighted mixed-effects models for repeated measures (MMRM), with weights derived from propensity-score models to balance pridopidine-treated participants and external controls from ENROLL-HD or TRACK-HD. Between-group differences in LS mean change were also estimated. To maintain adequate statistical power in the context of propensity-score weighting, sample trimming, and reduced effective sample sizes, particularly for smaller external cohorts, prespecified 90% confidence intervals (CI) were used for these externally controlled comparisons. For additional clinical interpretation, descriptive estimates of the percent slowing of decline were calculated at the Week 104 using percent slowing = *1 – (*Δ *pridopidine /* Δ *weighted natural history comparator)*, where Δ represents the LS mean change estimates obtained from MMRM trajectories for both the treated and comparator groups.

### Software

Propensity-score estimation, inverse-probability weighting, trimming, and MMRM analyses were implemented using SAS® 9.4, including logistic regression and PROC MIXED. Propensity-score specifications were aligned with previously published ENROLL-HD and TRACK-HD external-control applications in early HD ^14,30^.

## Results

### Participant Characteristics

Baseline demographic and disease characteristics are summarized in **Table 1** for the pridopidine cohort and for the ENROLL-HD and TRACK-HD cohorts before and after ATT propensity-score weighting with trimming (see **Methods**). A total of 90 participants who remained off-ADMs throughout the DBP and open-label extension contributed evaluable data through Week 104 and comprised the off-ADM analysis population (**Table 1**).

**Table 1.** Baseline characteristics of pridopidine-treated participants and propensity-score–weighted ENROLL-HD and TRACK-HD cohorts. Baseline demographic and clinical characteristics of participants who remained off-ADMs throughout the double-blind phase and entered the open-label extension, as compared with PS-weighted cohorts from ENROLL-HD and TRACK-HD. Characteristics are shown at the double-blind baseline. ATT propensity-score weighting with trimming was performed separately for each external cohort using demographic and disease-related covariates. Standardized mean differences (SMDs) are shown before and after weighting to assess covariate balance, with values closer to zero indicating improved balance.

| Outcome | Δ Decline<br>(ENROLL-HD) | Δ Decline<br>(Pridopidine) | P-values | % Slowing<br>(ENROLL-HD) | Δ Decline<br>(TRACK-HD) | Δ Decline<br>(Pridopidine) | P-values | %<br>Slowing<br>(TRACK-HD) |
| --- | --- | --- | --- | --- | --- | --- | --- | --- |
| cUHDRS | -1.59 | -0.65 | p<0.0001 | 59.1% | -1.81 | -0.66 | p<0.0001 | 63.5% |
| TFC | -1.43 | -0.50 | p<0.0001 | 65.0% | -1.08 | -0.41 | p=0.004 | 62.0% |
| SWR | -7.70 | -0.90 | p<0.0001 | 88.3% | -9.52 | -1.76 | p=0.0003 | 81.5% |
| SDMT | -1.57 | -0.95 | p=0.29 | 39.5% | -3.21 | -1.38 | p=0.06 | 57.0% |
| TMS | 6.39 | 3.62 | p=0.002 | 43.3% | 6.35 | 3.27 | p=0.02 | 48.5% |
| Q-Motor<br>FT-IOI | — | — | — | — | 88.22 | 19.59 | p<0.0001 | 77.8% |

Before weighting, external-cohort participants differed from the treated cohort with respect to age, baseline functional capacity, and selected cognitive and motor measures; after weighting, covariate balance improved across prespecified demographic and disease-related variables, including age, sex, CAG repeat length, baseline TFC, cUHDRS, SWR, SDMT, TMS, and geographic region in both external comparisons (see **Methods** and **Table 1**). Standardized mean differences (SMDs) were used to quantify covariate balance, with absolute values closer to 0 indicating better balance. Baseline disease severity in the treated cohort was consistent with early manifest HD, with preserved functional capacity and measurable cognitive and motor involvement.

### Two-Year Clinical Change as Compared with ENROLL-HD

Among participants receiving pridopidine treatment who remained off-ADMs, longitudinal changes in functional, composite, cognitive, and motor outcomes through Week 104 were compared with estimates from a PS–weighted ENROLL-HD cohort (**Figure 1**). All p-values reported for these external-comparator analyses are nominal and should be interpreted with caution. Additional details are reported in **Supplemental Table 2**.

**Figure 1.**
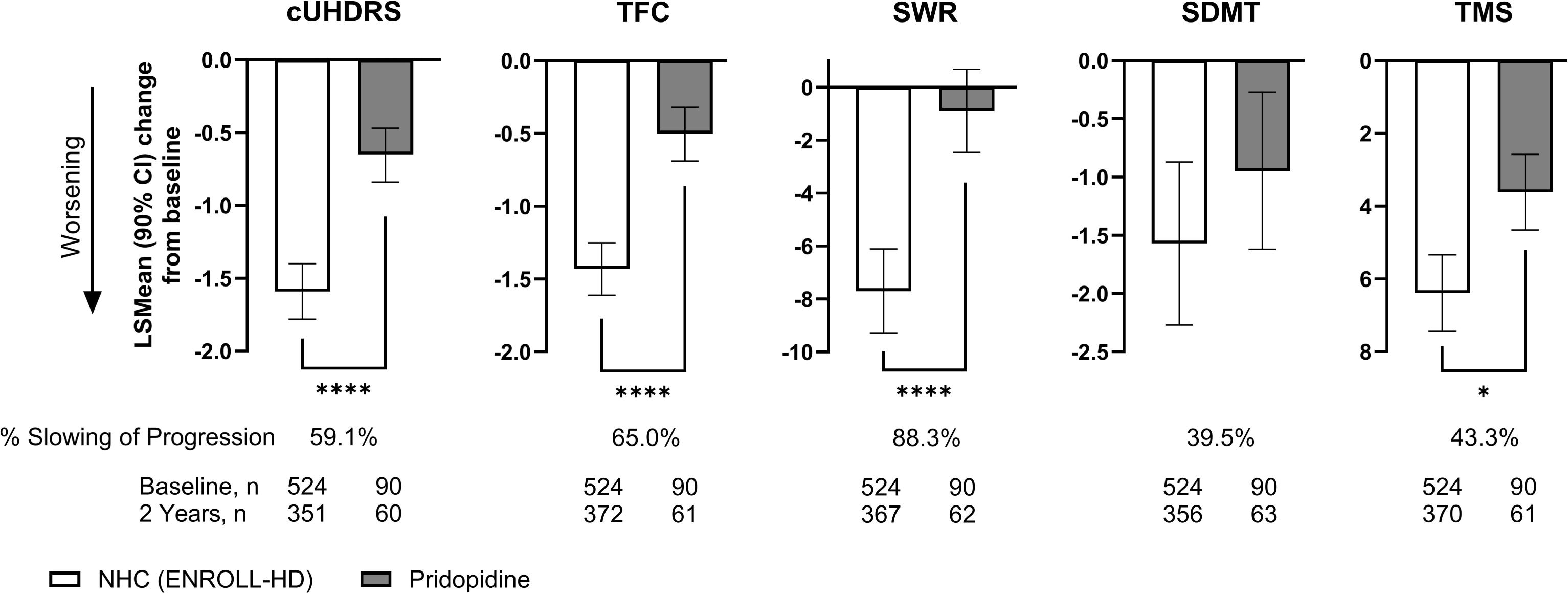
Outcomes in pridopidine-treated participants and propensity-score–weighted ENROLL-HD cohort. Data show change from double-blind baseline through Week 104 in composite (cUHDRS), functional (TFC), cognitive (SWR and SDMT), and motor (TMS) outcomes among participants receiving pridopidine treatment through week 104 who remained off-ADMs as compared with a PS–weighted cohort from ENROLL-HD. Negative values indicate worsening for cUHDRS, TFC, SWR, and SDMT; positive values indicate worsening for TMS, for which higher scores reflect greater motor impairment. Percent slowing was calculated relative to weighted natural-history estimates and for descriptive interpretation only. Bars represent LS mean change at Week 104 with 90% confidence intervals (CIs); * p<0.05; **** p<0.0001.

At Week 104, change from baseline in cUHDRS was more favorable in the continuous-treatment cohort than in the matched ENROLL-HD cohort (LS mean change −0.65 vs −1.59; between-group difference 0.94; 90% CI, 0.68 to 1.21; p<0.0001). A similar pattern was observed for functional capacity, with more favorable change in TFC in the treated cohort as compared with ENROLL-HD (−0.50 vs −1.43; between-group difference 0.93; 90% CI, 0.67 to 1.18; p<0.0001). Based on least-squares mean changes at Week 104, modeled estimates indicated 59.1% slowing of progression in cUHDRS and 65.0% slowing in TFC relative to ENROLL-HD (**Supplemental Table 1**). These modeled estimates are provided for descriptive interpretation only.

Cognitive outcomes showed differences favoring pridopidine in SWR and SDMT. Worsening in SWR was less among participants receiving pridopidine than among matched ENROLL-HD participants (−0.90 vs −7.70; between-group difference 6.80; 90% CI, 4.57 to 9.02; p<0.0001). For the SDMT, the treated cohort showed numerically less worsening than ENROLL-HD (−0.95 vs −1.57), with the confidence interval for the between-group difference including zero (difference 0.63; 90% CI, −0.35 to 1.60; p=0.29). Modeled estimates at Week 104 suggested 88.3% slowing of progression in SWR and 39.5% slowing in SDMT relative to ENROLL-HD (**Supplemental Table 1**), provided for descriptive interpretation only.

Motor progression, assessed by TMS (higher scores indicate worse motor function), showed a smaller increase (less worsening) in the treated cohort than in the external comparator (3.62 vs 6.39; between-group difference −2.77; 90% CI, −4.25 to −1.30; p=0.002). Modeled estimates at Week 104 suggested 43.3% slowing of TMS progression relative to ENROLL-HD (**Supplemental Table 1**), provided for descriptive interpretation only.

### Two-Year Clinical Change as Compared with TRACK-HD

Longitudinal changes through Week 104 among participants receiving pridopidine treatment and remaining off-ADMs were evaluated using TRACK-HD as a second PS–weighted external natural-history comparator (**Figure 2**). All p-values are nominal and should be interpreted with caution. Additional statistical details are reported in **Supplemental Table 1**.

**Figure 2.**
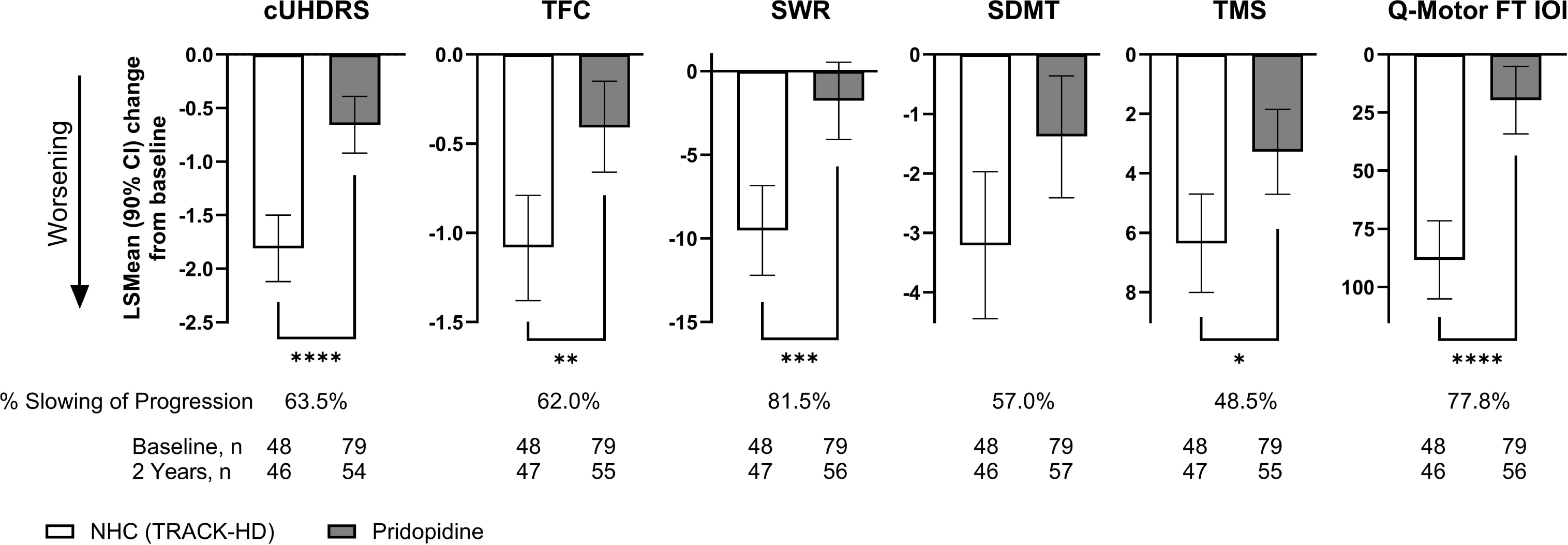
Outcomes in pridopidine-treated participants and propensity-score–weighted TRACK-HD cohort. Data show change from double-blind baseline through Week 104 in composite (cUHDRS), functional (TFC), cognitive (SWR and SDMT), and motor (TMS and Q-Motor FT-IOI) outcomes among participants receiving pridopidine treatment through week 104 who remained off-ADMs as compared with a PS–weighted cohort from TRACK-HD. Negative values indicate worsening for cUHDRS, TFC, SWR, and SDMT; positive values indicate worsening for TMS and Q-Motor FT-IOI, for which higher scores reflect worse performance. Percent slowing was calculated relative to weighted natural-history estimates and for descriptive interpretation only. Bars represent LS mean change at Week 104 with 90% confidence intervals (CIs); * p<0.05; ** p<0.01; *** p<0.001; **** p<0.0001.

At Week 104, cUHDRS decline showed less worsening among participants receiving pridopidine compared with the external cohort (−0.66 vs −1.81; between-group difference 1.15; 90% CI, 0.75 to 1.56; p<0.0001). Similarly, functional decline was smaller in the pridopidine treated cohort than in the external comparator, with a smaller reduction in TFC (LSMean change −0.41 vs −1.08; between-group difference 0.68; 90% CI, 0.29 to 1.06; p=0.004). Based on least-squares mean changes at Week 104, modeled estimates suggested 63.5% slowing of cUHDRS progression and 62.0% slowing of TFC progression relative to TRACK-HD (**Supplemental Table 1**). These modeled estimates are provided for descriptive interpretation only.

In cognitive outcome measures, SWR and SDMT showed differences favoring pridopidine. SWR worsening was less in the treated cohort than in the external TRACK-HD comparator (−1.76 vs −9.52; between-group difference 7.76; 90% CI, 4.23 to 11.29; p=0.0003). SDMT worsening was also numerically less as compared with the external cohort (−1.38 vs −3.21; between-group difference 1.82; 90% CI, 0.22 to 3.43; p=0.06). Descriptive estimates at Week 104 indicated 81.5% slowing of SWR progression and 57.0% slowing of SDMT progression relative to TRACK-HD (**Supplemental Table 1**).

TMS (higher scores indicate worse motor performance) increased to a lesser extent among participants receiving pridopidine than in the external cohort (3.27 vs 6.35; between-group difference −3.08; 90% CI, −5.26 to −0.89; p=0.02). Quantitative motor assessment using Q-Motor FT IOI showed less worsening in the treated cohort as compared with the external comparator (19.59 vs 88.22 ms; between-group difference −68.63 ms; 90% CI, −90.76 to −46.50; p<0.0001), showing less worsening of tapping interval time over two years. Relative to TRACK-HD, motor outcomes in the treated cohort progressed at a slower rate, corresponding to 48.5% slower progression on TMS and 77.8% slower progression on Q-Motor FT-IOI (**Supplemental Table 1**).

### Exploratory Outcomes in Participants Receiving Concomitant ADMs

To further explore whether less worsening with pridopidine treatment were also observed in the context of concomitant ADM use, we examined the primary and key secondary outcome measures from the main DBP, i.e., TFC and cUHDRS ^10^, in participants who completed 65 weeks of double-blind treatment, continued treatment through Week 104, and received recommended-dose ADMs at any time during the study. Given the exploratory nature of these analyses, and the absence of Type I error control, results are descriptive and should be interpreted cautiously.

Baseline demographic and clinical characteristics for these participants and their PS–weighted ENROLL-HD comparators are summarized in **Supplemental Table 2**. Dose cutoffs used to define recommended-dose ADM exposure are detailed in **Supplemental Table 3**. Sample sizes at Week 104 were limited, with 21 pridopidine-treated participants and 125 ENROLL-HD comparators contributing to the TFC analysis, and 19 and 115 participants, respectively, contributing to the cUHDRS analysis.

At Week 104, participants receiving pridopidine while on recommended dose ADMs had less worsening in both functional capacity and composite clinical status as compared with the matched ENROLL-HD cohort (**Figure 3**). Corresponding statistics are reported in **Supplemental Table 4.**

**Figure 3.**
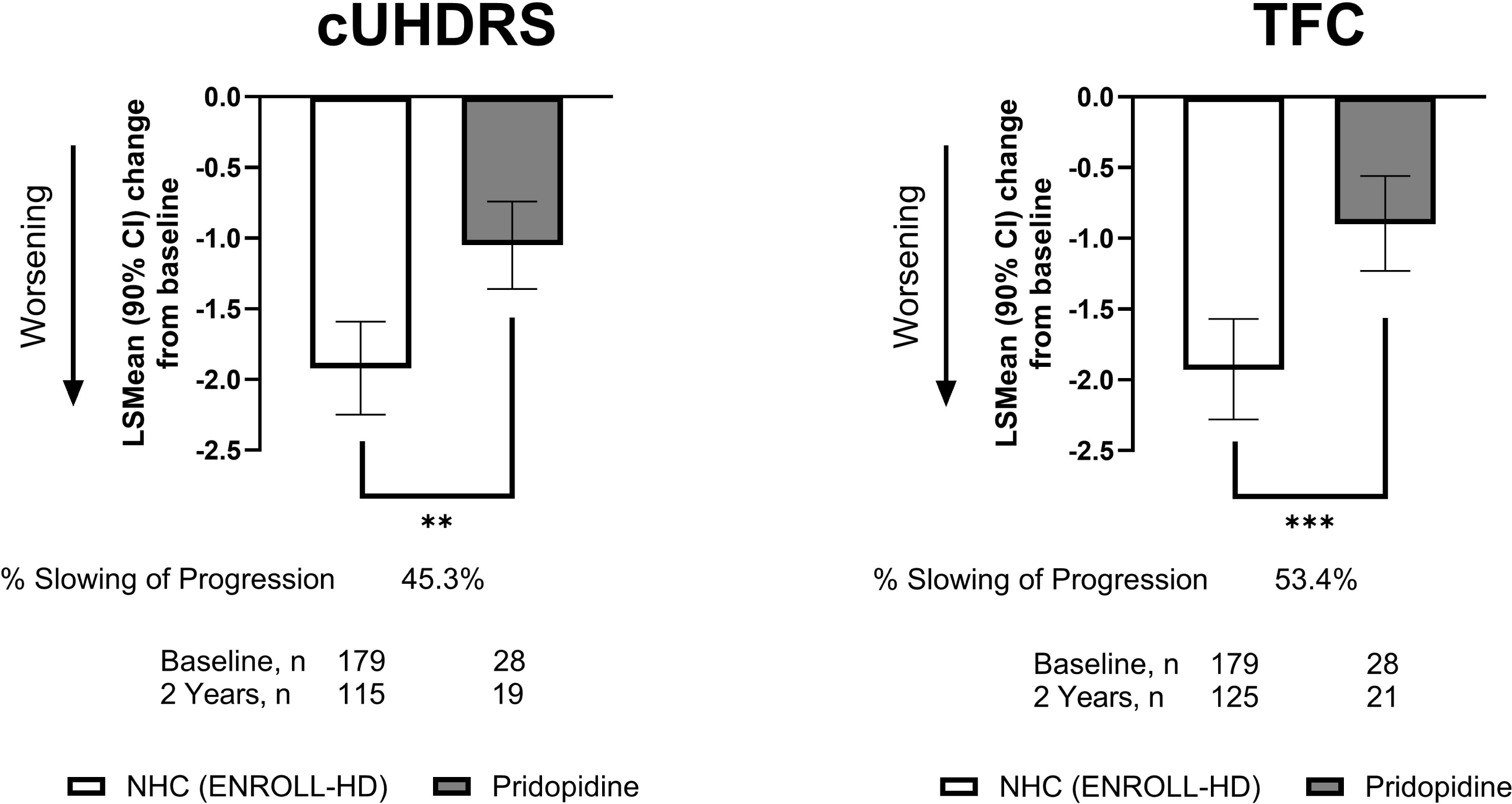
Outcomes in pridopidine-treated participants receiving recommended-dose antidopaminergic medications and propensity-score–weighted ENROLL-HD cohort. Exploratory data show change from double-blind baseline through Week 104 in functional (TFC) and composite (cUHDRS) outcomes among participants receiving pridopidine treatment through week 104 with concomitant recommended-dose antidopaminergic medications (ADMs) as compared with a PS–weighted cohort from ENROLL-HD. Negative values indicate worsening for both outcomes. Recommended-dose ADM exposure for tetrabenazine, deutetrabenazine, aripiprazole, and quetiapine was defined using dose cutoffs shown in **Supplemental Table 3**. Percent slowing was calculated relative to weighted natural-history estimates and for descriptive interpretation only. Bars represent LS mean change at Week 104 with 90% confidence intervals (CIs); ** p<0.01; *** p<0.001.

For TFC, the LS mean change from baseline at Week 104 was −0.90 among participants receiving pridopidine with recommended-dose ADMs, as compared with −1.93 in the propensity-score–weighted ENROLL-HD cohort, corresponding to a between-group difference of 1.03 (90% CI, 0.54 to 1.52; p<0.0006). For cUHDRS, the LS mean change was −1.05 with pridopidine and −1.92 in ENROLL-HD, yielding a between-group difference of 0.87 (90% CI, 0.42 to 1.33; p=0.0016).

When expressed descriptively, pridopidine in this exploratory analysis corresponded to 45.3% slowing of progression in cUHDRS and 53.4% slowing of progression in TFC as compared with the matched ENROLL-HD cohort at Week 104. Additional numerical results and model-derived estimates are provided in **Supplemental Table 4**. These descriptive differences were consistent with less worsening among participants receiving pridopidine with concomitant ADM exposure.

## DISCUSSION

The PROOF-HD OLE evaluated whether two-year disease trajectories under pridopidine treatment differed from those expected based on natural-history disease progression. As compared with PS–weighted comparators from two independent external cohorts (ENROLL-HD and TRACK-HD), participants receiving pridopidine exhibited slowing of progression through Week 104 across cUHDRS, TFC, SWR, and TMS, with quantitative Q-Motor findings in TRACK-HD. Together, these findings support sustained differences favoring pridopidine in observed outcomes over two years relative to expected early-HD progression.

ENROLL-HD and TRACK-HD are large, prospectively followed natural-history cohorts in HD that provide estimates of expected clinical progression in the absence of disease-modifying therapy ^6,12,31^. At Week 104, as compared with ENROLL-HD and TRACK-HD, between-group differences in LS mean change favored pridopidine treatment across cUHDRS, TFC, SWR, and TMS outcome measures, with the largest descriptive percent slowing observed for SWR (81.5–88.3% slowing as compared with both natural history cohorts). Similar differences were also observed in Q-Motor scores relative to TRACK-HD ^6^.

Our findings raise a clinically relevant question: Could the observed two-year differences under pridopidine treatment translate into preservation of function in early HD? Prior modeling and clinical-interpretation frameworks have suggested that a relatively modest 0.2–0.3 points per year reduction in the annual rate of progression, if sustained, may translate into clinically important delays in functional deterioration^7,32^.

In the present analysis, the cumulative differences across outcomes, approximately 0.9–1.2 points in cUHDRS and 0.7–1.0 points in TFC over two-years, were greater than would be expected based on untreated natural-history trajectories modeled from ENROLL-HD and TRACK-HD. When expressed relative to annualized projections, between-group differences of this magnitude exceed ranges proposed as clinically relevant for treatment-associated differences in early HD (Schobel et al., 2017).

Moreover, descriptive Week 104 estimates indicated ∼59–64% slowing of progression for cUHDRS and ∼62–65% for TFC as compared with expected untreated trajectories. Larger relative attenuation was observed for SWR (∼82–88%) and Q-Motor finger-tapping inter-onset interval (∼78%) (see **Supplementary Materials** for other outcomes). In particular, the Q-Motor finger-tapping assessment, due to its objective nature, has never exhibited placebo effects in previous randomized trials ^10,33^, supporting the interpretation of a sustained benefit of pridopidine. In this context and recognizing that statistical threshold effects cannot be formally confirmed in externally controlled analyses, the overall direction and consistency of differences observed across all outcomes, cUHDRS, TFC, SWR/SDMT, TMS, and Q-Motor FT-IOI, support the intuitive interpretation that sustained differences over two years with continuous pridopidine treatment may be viewed as “time saved” from experiencing disease progression ^7,13,32^. Given the open-label design and use of external comparators, these observations should be interpreted cautiously.

Because symptomatic therapies remain important in routine HD care, we conducted exploratory analyses evaluating pridopidine administered alongside recommended lower-dose VMAT2 inhibitors or select antipsychotics to approximate real-world treatment ^23,25,34^; while attempting to minimize confounding and outcome masking associated with potent D2 antagonism and CYP2D6-mediated interactions ^10,28,35^. Concomitant ADMs can complicate interpretation of longitudinal outcomes in HD, including by contributing to adverse effects that overlap with manifestations of disease progression; this challenge is acknowledged in regulatory labeling for ADMs ^24,30,34,36,37^. For example, tetrabenazine approved labeling notes that it may be difficult to distinguish adverse side effects from HD progression ^38^. Evidence from clinical and observational studies, as well as neuroimaging, suggests that sustained dopamine antagonism may impair cognitive and motor performance in a dose-dependent manner, which could attenuate or obscure changes measured by instruments such as cUHDRS ^15,21,24,27,34,39^. PROOF-HD predefined an off-ADM subgroup to reduce confounding ^10^. In agreement with this rationale, exploratory comparisons with PS-weighted ENROLL-HD participants showed differences favoring pridopidine in cUHDRS and TFC measures at Week 104 despite concomitant ADM exposure, although statistical inference is limited. Specifically, compared with PS–weighted ENROLL-HD participants, individuals receiving pridopidine and concomitant recommended dose ADMs demonstrated slower declines in cUHDRS and TFC through Week 104, corresponding to modeled estimates of 45.3–53.4% slowing of progression. These differences were nominally significant and of a magnitude that may be clinically meaningful when interpreted in the context of longitudinal disease progression. Although interpretation of these exploratory analyses remains limited by small sample size and the potential for residual confounding, between-group differences in progression remained observable under managed concomitant ADM regimens.

Long-term randomized, placebo-controlled follow-up in HD is constrained by the prolonged natural history of the disease and the practical difficulty of maintaining blinded placebo exposure over multiple years in a rare, progressive population ^1,3^. More broadly, real-world evidence analyses are inherently subject to measurement variability, incomplete covariate capture, and residual confounding. Thus, the presented findings should be interpreted cautiously given the inherent limitations of a single-arm open-label extension design. The extension did not include an internal placebo group, and long-term trajectories were therefore evaluated against PS-weighted external natural-history comparators. Although the external cohorts were balanced to the treated cohort using propensity-score methods, residual confounding from unmeasured differences could not be excluded, including differences in assessment schedules and procedures between the interventional trial and natural-history registries ^12,14^. In addition, we acknowledge that attrition may have also introduced selection effects and required additional missing-data assumptions ^40^. However, in this report, use of two independent natural-history cohorts (ENROLL-HD and TRACK-HD) provided an internal consistency check across datasets. Directionally consistent findings across multiple clinical outcomes, together with the observation of an effect on the objective Q-Motor assessment support an interpretation of sustained benefits in observed disease trajectories with pridopidine treatment.

In conclusion, over two years of follow-up, continuous pridopidine treatment was associated with slowing of progression based on PS-weighted external natural-history cohorts across functional, cognitive, motor, and quantitative motor assessment in early HD. Although the open-label design and use of external comparators do not establish causality, consistent findings across outcomes and two independent external cohorts support an interpretation of sustained slowing of progression over two years, most directly framed as time preserved before reaching clinically meaningful disease milestones.

## Data Availability

De-identified participant data (IDP) with data dictionaries, along with key documents, the protocol, SAP, and informed-consent template, for the parent PROOF-HD study and its open-label extension will be available to qualified researchers starting six months after publication and for five years thereafter.
Access is limited to investigators at academic or non-profit institutions pursuing scientifically sound, ethics-approved analyses that align with the scientific objectives of the PROOF-HD program or address closely related questions. Because of ethical and legal constraints on participant privacy (e.g., GDPR), data are not placed in a public repository. Redacted versions of the PROOF-HD protocol and SAP can be accessed here: https://ghi-muenster.de/protocols/proof-hd and https://ghi-muenster.de/protocols/proof-hd-sap.
To request access, contact the Sponsor at. Requests are reviewed by the Sponsor or its data access committee, with a decision provided within 90 days. Approved users enter a Data Use Agreement that prohibits re-identification, disallows unauthorized data sharing, and limits use to the agreed purposes. Authorship and acknowledgments follow the International Committee of Medical Journal Editors (ICMJE) Recommendations (2025). A DUA template is available on request. No third-party proprietary datasets were used; all data were collected and analyzed by the investigators and the Sponsor as described in Methods.

https://ghi-muenster.de/protocols/proof-hd

https://ghi-muenster.de/protocols/proof-hd-sap

## Acknowledgements

The study was sponsored and funded by Prilenia Therapeutics, which supported the study design, trial oversight and conduct, site operations, data collection, statistical analysis, and manuscript development. The sponsor had final responsibility for verifying the accuracy of the data. We thank the participants in PROOF-HD and their families and caregivers. We acknowledge the contributions of site staff, sponsor teams, and the Huntington Study Group (HSG^®^) for coordination and support. We especially recognize the site principal investigators and study coordinators whose efforts were essential to the parent trial execution.

## Disclosure

Reilmann: founder/owner of GHI and QuantiMedis; consulting/clinical operations/Q-Motor services for Prilenia; European PI/coordinating investigator (PROOF-HD). Feigin: research support to NYU from Prilenia for PROOF-HD (North America PI). Rosser: European co-PI in Prilenia-sponsored trials with payments to Cardiff University. Kostyk: site investigator (PROOF-HD). Other authors report no competing interests. Anderson: scientific advisor to Prilenia; site investigator (PROOF-HD). Tan, Chen, Hand, Geva: employees of Prilenia (stock options). Hayden: founder, CEO of Prilenia; holds equity.

## Author Contributions

**Ralf Reilmann**

Conceived and designed the study; served as European Principal Investigator; led clinical oversight and coordination in PROOF-HD (Europe).

**Andrew M. Tan**

Contributed to clinical data analysis and interpretation; executed manuscript development and supervised its final production and submission with all authors.

**Anne E. Rosser**

Conceived and designed the study; served as European co-lead; led clinical oversight and coordination in PROOF-HD (Europe).

**Kelly Chen**

Contributed statistical analysis and methodology, clinical data analysis and interpretation.

**Andrew Feigin**

Served as the North American Principal Investigator; led clinical oversight and coordination in PROOF-HD (North America).

**Karen Elta Anderson**

Contributed clinical interpretation, insight, and manuscript draft and editorial support.

**Sandra K. Kostyk**

Served as North American co-lead; led clinical oversight and coordination in PROOF-HD (North America). Contributed clinical interpretation, insight, and editorial support.

**Randal Hand**

Contributed to clinical interpretation, data visualization, and provided editorial support.

**Michal Geva**

Conceived and designed the study; contributed to clinical data analysis and interpretation; editorial support in manuscript development; involved in funding acquisition and study sponsorship.

**Michael R. Hayden**

Conceived and designed the study; contributed to clinical data analysis and interpretation; involved in funding acquisition and study sponsorship. Supervised study and manuscript development.

## Data Availability

De-identified participant data (IDP) with data dictionaries, along with key documents, the protocol, SAP, and informed-consent template, for the parent PROOF-HD study and its open-label extension will be available to qualified researchers starting six months after publication and for five years thereafter.

Access is limited to investigators at academic or non-profit institutions pursuing scientifically sound, ethics-approved analyses that align with the scientific objectives of the PROOF-HD program or address closely related questions. Because of ethical and legal constraints on participant privacy (e.g., GDPR), data are not placed in a public repository. Redacted versions of the PROOF-HD protocol and SAP can be accessed here: https://ghi-muenster.de/protocols/proof-hd and https://ghi-muenster.de/protocols/proof-hd-sap.

To request access, contact the Sponsor at. Requests are reviewed by the Sponsor or its data access committee, with a decision provided within 90 days.

Approved users enter a Data Use Agreement that prohibits re-identification, disallows unauthorized data sharing, and limits use to the agreed purposes. Authorship and acknowledgments follow the International Committee of Medical Journal Editors (ICMJE) Recommendations (2025). A DUA template is available on request. No third-party proprietary datasets were used; all data were collected and analyzed by the investigators and the Sponsor as described in Methods.

---

**Supplemental Table 1.** Estimated change and percent slowing of outcomes in pridopidine-treated participants and propensity-score–weighted ENROLL-HD and TRACK-HD cohorts. Estimated change from double-blind baseline to Week 104 for functional, composite, cognitive, and motor outcomes among participants receiving pridopidine treatment who remained off-ADMs as compared with PS–weighted cohorts from ENROLL-HD and TRACK-HD. Change (Δ) values represent model-derived LS mean changes. Percent slowing was calculated relative to weighted natural-history estimates from each external cohort. No adjustment for multiplicity was performed; therefore, p values are shown for nominal, descriptive comparison only. Q-Motor FT-IOI was available only for the TRACK-HD comparison because this measure is not collected in ENROLL-HD.

| Characteristic | Pridopidine Week 65 Completers<br>(Lower/Recommended ADM Dose) | ENROLL-HD PS-Matched | Std Mean Diff (SMD)<br>vs ENROLL-HD<br>(before PS- match) | Std Mean Diff (SMD)<br>vs ENROLL-HD<br>(after PS- match) |
| --- | --- | --- | --- | --- |
| N analyzed | 28 | 179 |  |  |
| Age, years | 56.3 (11.44) | 57.5 (13.50) | 0.07 | -0.11 |
| Sex, % female | 60.0 | 70.0 | 0.08 | -0.10 |
| Region, % EU | 60.0 | 60.0 | -0.15 | -0.05 |
| Education, % above HS | 70.0 | 70.0 | 0.40 | 0.05 |
| CAG repeat length | 43.5 (4.02) | 43.1 (3.69) | -0.02 | 0.09 |
| CAP / Disease Burden Score | 110.6 (11.84) | 110.7 (17.50) | -0.01 | 0.00 |
| TFC | 9.7 (1.82) | 9.7 (1.72) | 0.17 | -0.01 |
| cUHDRS | 8.5 (2.31) | 8.4 (2.20) | 0.14 | 0.06 |
| SDMT | 21.5 (8.50) | 21.1 (8.24) | -0.10 | 0.05 |
| SWR | 58.7 (17.25) | 58.0 (16.03) | 0.07 | 0.04 |
| TMS | 31.9 (9.20) | 32.9 (10.46) | -0.26 | -0.10 |

**Supplemental Table 2.**
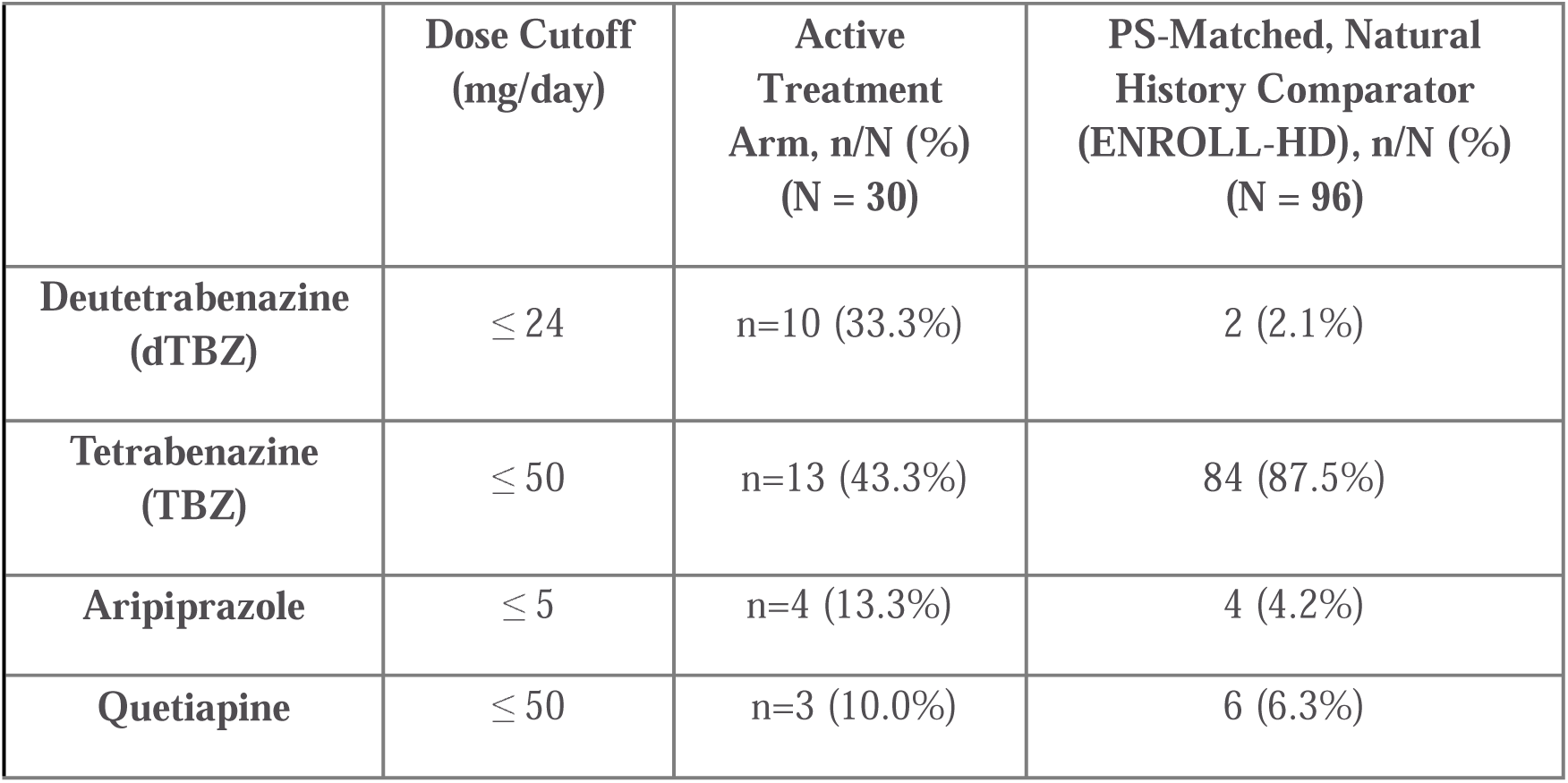
Baseline characteristics of pridopidine-treated participants receiving recommended-dose antidopaminergic medications and propensity-score–weighted ENROLL-HD cohort. Baseline demographic and clinical characteristics of participants receiving pridopidine treatment with concomitant recommended-dose ADMs as compared with ENROLL-HD. Characteristics are shown at the double-blind baseline. Recommended-dose ADM exposure was defined using dose cutoffs shown in **Supplemental Table 3**. External cohort participants were selected using PS methods based on prespecified demographic and disease-related covariates. Standardized mean differences (SMDs) are shown before and after ATT PS-weighting with trimming to assess covariate balance.

**Supplemental Table 3.** Dose cutoff values for antidopaminergic medications used in analyses. Dose cutoff values reflect label recommended-dose antidopaminergic medication exposure in exploratory analyses, together with the number and percentage of participants meeting these criteria in the active treatment cohort and the PS–weighted ENROLL-HD comparator cohort. Participants receiving more than one ADM concurrently were excluded. Antidopaminergic medication categories included VMAT2 inhibitors, i.e., deutetrabenazine and tetrabenazine, and antipsychotic drugs, i.e., aripiprazole and quetiapine. Percentages were calculated using the total cohort values shown in the column headers.

| **Outcome**<br>**(Week 65 completers)** | **Δ Decline**<br>**(ENROLL-HD)**<br>**(n-value)** | **Δ Decline**<br>**(Pridopidine + ADM)**<br>**(n-value)** | **P-values** | **% Slowing**<br>**(ENROLL-HD)** |
| --- | --- | --- | --- | --- |
| **cUHDRS** | −0.91 (n = 115) | −0.20 (n = 19) | 0.0051 | 45.3% |
| **TFC** | −1.29 (n = 125) | −0.17 (n = 21) | <0.0001 | 53.4% |

**Supplemental Table 4.**
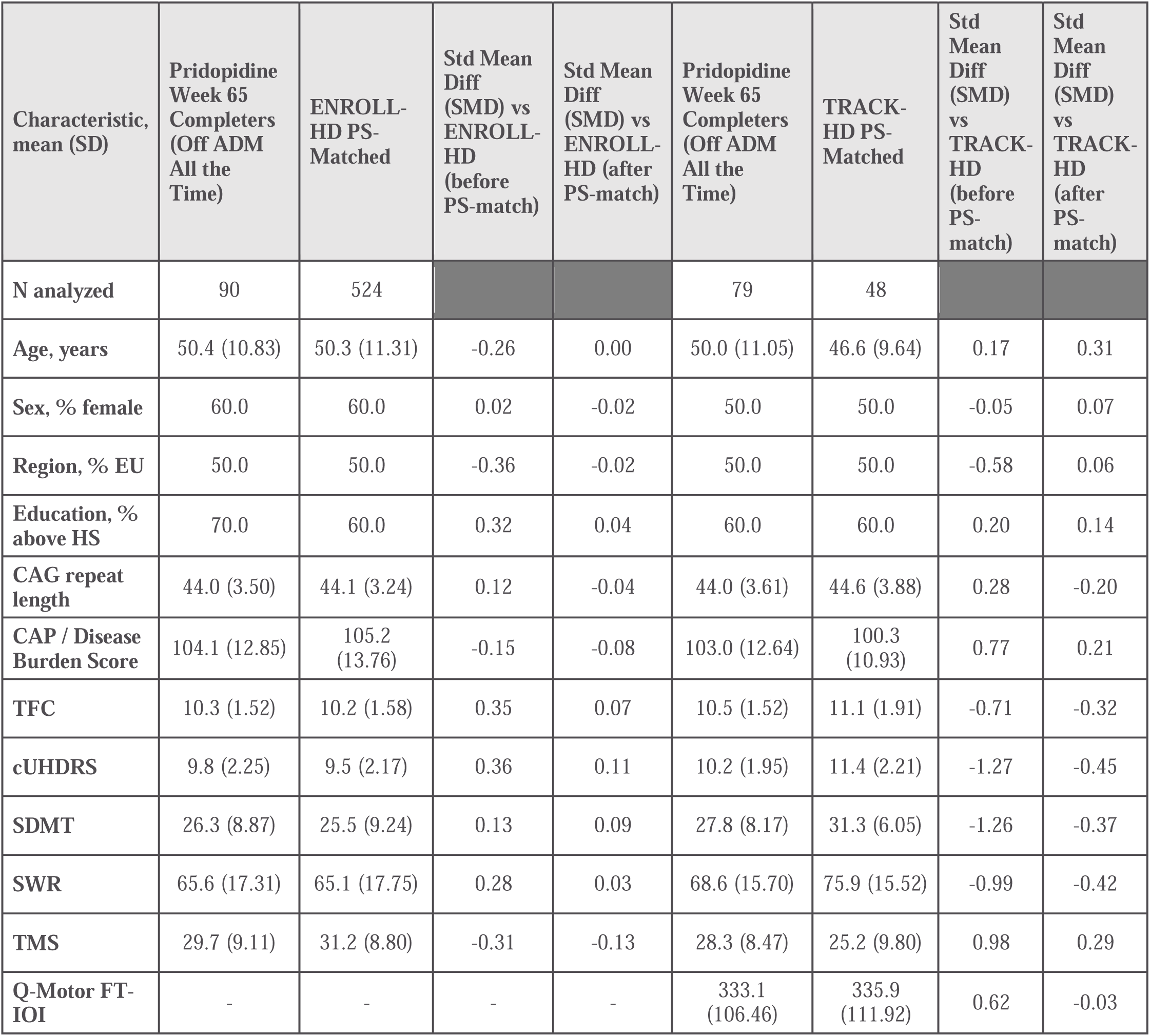
Estimated change and percent slowing of outcomes in pridopidine-treated participants receiving recommended-dose antidopaminergic medications and propensity-score–weighted ENROLL-HD cohort. Estimated change from double-blind baseline to Week 104 for functional and composite outcomes among participants receiving pridopidine treatment with concomitant recommended-dose ADMs as compared with a PS–weighted ENROLL-HD cohort. Change (Δ) values represent model-derived LS mean changes. Percent slowing was calculated relative to weighted natural-history estimates from ENROLL-HD. No adjustment for multiplicity was performed; therefore, p values are shown for nominal, descriptive comparison only. Recommended-dose ADM exposure was defined using dose cutoffs shown in **Supplemental Table 3**.

| Characteristic,<br>mean (SD) | Pridopidine<br>Week 65<br>Completers<br>(Off ADM<br>All the<br>Time) | ENROLL-HD PS-Matched | Std Mean<br>Diff<br>(SMD) vs<br>ENROLL-HD<br>(before<br>PS-match) | Std Mean<br>Diff<br>(SMD) vs<br>ENROLL-HD (after<br>PS-match) | Pridopidine<br>Week 65<br>Completers<br>(Off ADM<br>All the<br>Time) | TRACK-HD PS-Matched | Std<br>Mean<br>Diff<br>(SMD)<br>vs<br>TRACK-HD<br>(before<br>PS-match) | Std<br>Mean<br>Diff<br>(SMD)<br>vs<br>TRACK-HD<br>(after<br>PS-match) |
| --- | --- | --- | --- | --- | --- | --- | --- | --- |
| N analyzed | 90 | 524 |  |  | 79 | 48 |  |  |
| Age, years | 50.4 (10.83) | 50.3 (11.31) | -0.26 | 0.00 | 50.0 (11.05) | 46.6 (9.64) | 0.17 | 0.31 |
| Sex, % female | 60.0 | 60.0 | 0.02 | -0.02 | 50.0 | 50.0 | -0.05 | 0.07 |
| Region, % EU | 50.0 | 50.0 | -0.36 | -0.02 | 50.0 | 50.0 | -0.58 | 0.06 |
| Education, %<br>above HS | 70.0 | 60.0 | 0.32 | 0.04 | 60.0 | 60.0 | 0.20 | 0.14 |
| CAG repeat<br>length | 44.0 (3.50) | 44.1 (3.24) | 0.12 | -0.04 | 44.0 (3.61) | 44.6 (3.88) | 0.28 | -0.20 |
| CAP / Disease<br>Burden Score | 104.1 (12.85) | 105.2<br>(13.76) | -0.15 | -0.08 | 103.0 (12.64) | 100.3<br>(10.93) | 0.77 | 0.21 |
| TFC | 10.3 (1.52) | 10.2 (1.58) | 0.35 | 0.07 | 10.5 (1.52) | 11.1 (1.91) | -0.71 | -0.32 |
| cUHDRS | 9.8 (2.25) | 9.5 (2.17) | 0.36 | 0.11 | 10.2 (1.95) | 11.4 (2.21) | -1.27 | -0.45 |
| SDMT | 26.3 (8.87) | 25.5 (9.24) | 0.13 | 0.09 | 27.8 (8.17) | 31.3 (6.05) | -1.26 | -0.37 |
| SWR | 65.6 (17.31) | 65.1 (17.75) | 0.28 | 0.03 | 68.6 (15.70) | 75.9 (15.52) | -0.99 | -0.42 |
| TMS | 29.7 (9.11) | 31.2 (8.80) | -0.31 | -0.13 | 28.3 (8.47) | 25.2 (9.80) | 0.98 | 0.29 |
| Q-Motor FT-IOI | - | - | - | - | 333.1<br>(106.46) | 335.9<br>(111.92) | 0.62 | -0.03 |

## Notes

### Clinical Trial

NCT04556656

### Author Declarations

Ethics committee/IRB of the University of Rochester Medical Center gave ethical approval for this work

