## Supplemental Methods for "Long-Term Slowing of Progression in Huntington’s Disease with Pridopidine Treatment"

### Table of Contents (Supplemental Appendix)

#### Supplementary Methods

##### ****Study Design****

PROOF-HD was a Phase 3, randomized, double-blind, placebo-controlled study followed by a prespecified open label extension (NCT04556656). In the double-blind period (DBP), the primary endpoint was assessed at Week 65. After completing the DBP, participants could enter a planned open-label extension (OLE) to evaluate long-term outcomes under continuous pridopidine exposure. For analyses presented here, outcomes were evaluated from the randomized double-blind phase (DBP) baseline through Week 104. All changes from baseline were calculated relative to the DBP baseline.

All participants entering the follow-up received open-label pridopidine 45 mg twice daily. Although all patients received active treatment in the extension, the randomized treatment assignments from the DBP remained undisclosed to investigators, participants, and sponsor personnel throughout the follow-up period. The blind was maintained until the open-label extension database lock, finalized on May 24, 2024.

A concurrent, long-term placebo group was not feasible. Accordingly, the statistical analysis plan (SAP) prespecified the use of natural-history external comparator cohorts for contextualizing long-term clinical progression. Comparator groups were constructed from the ENROLL-HD and TRACK-HD observational studies through propensity score-based weighting methodologies, as detailed in the SAP. Follow-up visits for comparators were aligned through visit-windows corresponding to nominal Year 1 through, Year 2, and Year 3 visits to ensure comparable durations.

Two implemented SAPs addenda defined the Off-ADM populations used for the long-term analyses. SAP Addendum 1.0 (January 18, 2024) specified a primary focus on OLE participants not receiving antidopaminergic medications (Off-ADM), given evidence that such medications could obscure treatment-associated effects and were associated with accelerated progression. This report uses the broader, more inclusive Off-ADM cohort defined at the double-blind baseline and followed through Week 104 as the primary analysis population.

The protocol and all amendments were approved by institutional review boards or independent ethics committees at each participating site. The trial was conducted according to the principles of the Declaration of Helsinki and the International Council for Harmonisation Good Clinical Practice guidelines. Written informed consent was obtained from all participants at initial enrollment, with reconsent required for participation in the long-term follow-up.

##### ****Participants****

###### ****Eligibility for Long-Term Continuous-Treatment Analysis****

Participants eligible for inclusion in the long-term continuous-treatment analysis were those who completed the End-of-Study visit of the double-blind period (DBP) without major protocol deviations and who proceeded into the open-label extension. All participants initiating the extension entered a two-week titration phase of pridopidine 45 mg once daily before receiving the open-label target dose of 45 mg twice daily.

To minimize confounding arising from concomitant medications, the primary analytic population for DBP portion was restricted to participants who were Off ADMs during this period. For the open-label extension, the analytic population included participants who remained off ADMs throughout the DBP and extension and contributed evaluable follow-up data through Week 104.

###### ****ADM Exposure Classification****

Because ADMs, including VMAT2 inhibitors and dopamine D₂ receptor–acting neuroleptics, may confound pridopidine-associated effects or accelerate clinical decline, ADM exposure was explicitly classified. The Off-ADM group consisted of participants who did not receive any ADMs at any time during the DBP or the extension.

A secondary subgroup consisted of participants who were Off-ADM or receiving recommended doses of selected ADMs. Based on the tabulated SAP definitions, the low-dose thresholds were:

- Tetrabenazine ≤ 50 mg/day
- Deutetrabenazine ≤24 mg/day
- Aripiprazole ≤ 5 mg/day
- Quetiapine ≤ 50 mg/day

Participants receiving more than one ADM simultaneously or any dose of olanzapine or risperidone were excluded because these agents exhibit high-affinity D₂ antagonism and dose-dependent receptor occupancy known to confound motor and cognitive outcomes^1–5.^

These ADM-defined subgroups were used as exploratory populations to assess whether long-term pridopidine treatment demonstrated directionally consistent trajectories in the presence of low-dose symptomatic therapy. Exploratory ADM analyses in this report focus on tetrabenazine, deutetrabenazine, aripiprazole, and quetiapine.

**Analysis Populations**

Efficacy analyses were performed within the Off-ADM pridopidine continuous-treatment populations. The primary analysis population consisted of the modified intent-to-treat (mITT) cohort restricted to Off-ADM participants during the DBP. For the OLE portion, only participants who entered the OLE and remained off ADMs were included. This cohort formed the basis for propensity-score model fitting and all external-comparator analyses at Week 104.

An additional exploratory population consisted of participants receiving lower recommended doses of tetrabenazine, deutetrabenazine, aripiprazole, or quetiapine, consistent with predefined thresholds (see **Supplemental Table 3**). Analyses in this subgroup were reported, descriptively, using the same mixed-model framework as the primary analyses.

##### ****External Comparator Cohorts****

###### ****Cohort Selection****

As a long-term placebo group was not feasible, efficacy analyses for the follow-up relied on external comparator cohorts drawn from two established longitudinal observational studies: ENROLL-HD and TRACK-HD. ENROLL-HD is a large, multinational, prospective natural-history registry that includes more than 20,000 participants across 23 countries and provides annual assessments of functional, cognitive, and motor outcomes using instruments identical to those in PROOF-HD. TRACK-HD is a deeply phenotype, multicenter biomarker study that evaluated individuals with premanifest or early Huntington’s disease through annual follow-up. Both studies collected the core outcomes required for comparison with PROOF-HD, including Total Functional Capacity (TFC), the composite Unified Huntington’s Disease Rating Scale (cUHDRS), Stroop Word Reading (SWR), Symbol Digit Modalities Test (SDMT), and Total Motor Score (TMS). TRACK-HD additionally provided Quantitative Motor (Q-Motor) assessments—specifically the Finger-Tapping Inter-Onset Interval (FT-IOI)—which were not available in ENROLL-HD and allowed for comparator analyses of objective motor function.

###### ****Alignment With PROOF-HD****

External cohort follow-up periods were aligned to the PROOF-HD double-blind baseline and mapped to the nearest comparable annual visit intervals in ENROLL-HD and TRACK-HD. Because both observational studies used annual assessment schedules, PROOF-HD follow-up to 52 and 104 weeks corresponded to Year 1 and Year 2 visits in the external cohorts. Visits in ENROLL-HD and TRACK-HD were assigned to Year 1, Year 2 analysis time points using prespecified day windows centered on approximately 12 and 24 months after baseline. These windows were applied consistently across analyses to ensure comparable exposure durations between PROOF-HD and the external comparators. For Q-Motor outcomes, only TRACK-HD contributed data because ENROLL-HD did not collect Q-Motor assessments.

###### ****External Comparator Inclusion Criteria****

Comparator participants were selected to match the eligibility characteristics of the PROOF-HD follow-up population as closely as possible. For ENROLL-HD, inclusion criteria required age ≥25 years, a pathogenic CAG repeat length ≥36, Diagnostic Confidence Level (DCL) of 4, functional capacity of TFC ≥7 (corresponding to early manifest stages 1–2), TMS ≥20, Independence Scale ≤90, and residence in North America or Europe. TRACK-HD participants were required to have a clinical motor diagnosis consistent with manifest Huntington’s disease. Because the primary PROOF-HD analyses focused on participants Off ADMs, external comparator subjects were likewise required to be Off ADMs during the relevant assessment periods.

Propensity score weighting (PSW) served as the primary method for balancing external controls to the PROOF-HD analysis population. Covariates included in the propensity models were age, education level (high school or below, above high school), sex (female, male), region (North America, Europe), CAG repeat length, baseline CAP100, baseline TFC, baseline TMS, baseline SWR, and baseline SDMT.

###### ****Handling of Outcome Availability Differences****

Differences in assessment schedules between the observational cohorts required outcome-specific decisions regarding comparator selection. TRACK-HD provided Q-Motor FT-IOI assessments, enabling comparison with PROOF-HD on objective motor outcomes; ENROLL-HD did not include Q-Motor and therefore could not contribute to those analyses. All analyses were conducted with outcome-appropriate comparator cohorts to maintain consistency of available measures and minimize imputation requirements.

###### ****Clinical Outcomes****

Efficacy outcomes for the follow-up period were prespecified and aligned with the core clinical measures evaluated during the double-blind phase. Functional progression was assessed using the Total Functional Capacity (TFC) score. Global clinical progression was evaluated with the composite Unified Huntington’s Disease Rating Scale (cUHDRS), which integrates TFC, Total Motor Score (TMS), Stroop Word Reading (SWR), and the Symbol Digit Modalities Test (SDMT). Cognitive performance was evaluated with SWR and SDMT, and motor function was assessed using TMS. Objective motor performance was measured through the Q-Motor Finger Tapping Inter-Onset Interval (FT-IOI) mean, derived from the TRACK-HD dataset, which served as the sole source of external comparator data for Q-Motor FT-IOI.

###### ****Timepoints****

Analyses focused on change from the DBP baseline to Week 104, representing two years of continuous pridopidine treatment. PROOF-HD included scheduled assessments of change from DBP baseline at Week 26, 39, 52, 65, 78, and 104 (OLE Week 26), whereas external comparator cohorts provided data at Week 52 and 104. Treatment differences versus external controls were estimated using all available scheduled follow-up visits within the mixed-model repeated-measures framework. No analyses used OLE baselines, maintaining consistency with the continuous-treatment framework. Q-Motor outcomes were evaluable only at annual TRACK-HD visits.

##### ****Interpretation Framework****

Efficacy outcomes were summarized using least-squares (LS) mean change from baseline estimated from weighted mixed-effects models for repeated measures (MMRM), with weights derived from propensity-score models to balance pridopidine-treated participants and external controls from ENROLL-HD or TRACK-HD. Between-group differences in LS mean change were also estimated, enabling interpretation of long-term treated trajectories relative to matched natural-history cohorts. For additional clinical interpretability, descriptive estimates of percent slowing of decline were calculated by comparing annualized progression rates in the treated cohort with corresponding rates from the PS–weighted natural-history trajectories.

To maintain adequate statistical power in the context of propensity-score weighting, sample trimming, and reduced effective sample sizes, particularly for smaller external cohorts, prespecified 90% confidence intervals (CI) were used for these externally controlled comparisons. For additional clinical interpretation, descriptive estimates of the percent slowing of decline were calculated at the Week 104 using percent slowing = *1 – (Δ pridopidine / Δ weighted NHC)*, where Δ represents the LS mean change estimates obtained from MMRM trajectories for both the treated and comparator groups.

**Statistical Analysis**

###### ****Propensity Score Matching with Natural History Cohorts****

Separate propensity-score (PS) models were developed for ENROLL-HD and TRACK-HD under an average-treatment-effect-on-the-treated (ATT) framework, using the PROOF-HD primary Off-ADM analysis populations for DBP and OLE as the treated group. Logistic regression was used to estimate the PS. Baseline covariates in each model included age, sex, CAG repeat length, TFC, cUHDRS, SWR, SDMT, TMS, and geographic region. Additional ENROLL-HD–specific covariates included education level. CAP100, as an interaction term between age and CAG, was included where available to align with natural-history model specifications.

###### ****Weighting, Trimming, and Balance Assessment****

ATT weights under inverse-probability-of-treatment weighting were applied to participants in the natural-history cohorts. Trimming thresholds prespecified in the SAP excluded individuals with PS <0.05 or >0.95 to reduce extrapolation beyond the zone of empirical overlap. Standardized mean differences (SMDs) were used to assess covariate balance before and after weighting, targeting ≤0.1 and ideally ≤0.05 for major prognostic variables.

Mixed-model repeated-measures (MMRM) analyses with ATT weights from the above PS model were used to estimate least-squares (LS) mean change from baseline through Week 104. Fixed effects included visit, baseline value of the outcome, HD stage, and region, with a random intercept for each participant. Residual covariance structure followed the hierarchy specified in the SAP. An unstructured covariance matrix was used when convergence permitted, with sequential fallback to Toeplitz with heterogeneity, ARH(1), or ARMA(1,1). No imputation was performed; the model assumes data are missing at random. The MMRM model structure used for exploratory ADM analyses was identical but was applied to the relevant pridopidine-treated subgroups (Off-ADM and recommended-dose ADM) and, where external comparators were constructed, incorporated the same PS-weighting scheme as in the primary analyses.

##### ****Exploratory analyses of ADM combinations with pridopidine****

Exploratory analyses evaluated clinical trajectories among participants receiving pridopidine together with lower recommended doses of tetrabenazine, deutetrabenazine, aripiprazole, or quetiapine (see **Supplemental Table 3**). These analyses summarized LSmean change from baseline within each ADM-defined subgroup and, in selected models, compared Week-104 outcomes with PS-weighted natural-history cohorts constructed using the same covariates and ATT framework as in the primary analyses. All ADM combination analyses were SAP specified as exploratory and non-confirmatory.

###### ****Software****

Propensity-score estimation, inverse-probability weighting, trimming procedures, and the mixed-model repeated-measures analyses were implemented using SAS^®^ 9.4, including logistic regression, PROC MIXED and PROC PSMATCH. Construction of the external comparator cohorts followed the analytic frameworks defined in the ENROLL-HD and TRACK-HD statistical analysis plans, including cohort selection rules, longitudinal visit mapping, and handling of missingness. All analyses were conducted on locked, version-controlled datasets, and PS specifications were aligned with previously published ENROLL-HD and TRACK-HD external-control applications in early HD ^6,7^.

#### Supplemental Results

##### Participant Characteristics

Baseline characteristics for participants receiving continuous pridopidine treatment together with recommended-dose antidopaminergic medications are presented in **Supplemental Table 2**, with corresponding exposure frequencies summarized in **Supplemental Table 3**. These participants exhibited baseline functional, cognitive, and motor profiles consistent with early manifest disease, and post-weighting balance with ENROLL-HD comparators was improved across major demographic and disease-related variables. Note that **Supplemental Tables 1–4** summarize **related but non-identical analysis sets** (baseline/balance, exposure-frequency summaries, and outcome-specific model samples), denominators differ by table and by outcome.
